# Intensive care unit (ICU) healthcare workers’ experiences during the COVID-19 pandemic in Chile: A qualitative interview study

**DOI:** 10.1101/2025.09.28.25336848

**Authors:** Ana Castro-Avila, Javiera Aguilera

## Abstract

**Aim:** To explore how intensive care unit healthcare workers experienced their hospital response prior to and during the COVID-19 pandemic in seven centres in Chile.

**Methods:** We conducted semi-structured telephone interviews with ICU consultants, nurses, physiotherapists and healthcare assistants in seven hospitals in Chile. Healthcare professionals enrolled through QR codes displayed on posters in their staff rooms. The interviews were recorded, transcribed verbatim and analysed following the principles of framework analysis.

**Findings:** We interviewed 38 healthcare professionals between November 2020 and March 2022, of which 15(39%) were physiotherapists, 9(24%) were nurses, 7(18%) were healthcare assistants, of whom 21(56%) had worked in ICU for three to 10 years. We identified seven themes: 1) Units opened and converted to ICU “as the pandemic unfolded”; 2) “change of usual roles” where more experienced professionals took on provisional managerial roles alongside their clinical duties; 3) “social isolation, exhaustion, and lack of psychological support”; 4) “Fear of becoming a vector for infection”; 5) “Keep calm and carry on” mentality where professionals kept working regardless of their own wellbeing; 6) “Depersonalisation”; and 7) “Uncertainty” of the best course of treatment.

**Conclusions:** Our interviewees perceived the institutional preparation as insufficient, which led to a more-than-expected workload increase, exhaustion, and personal sacrifices to maintain standards of care. Institutional measures to support the workforce were perceived as late, inadequate, or insufficient.

**Implications for Clinical Practice:** ICU workers are a scarce and valuable resource during public health emergencies, planning should be in place to rapidly increase ICU staff in the event of a new prolonged emergency without burning out existing human resources. Appropriate psychological and emotional support should be provided in a timely fashion.

## Introduction

The COVID-19 pandemic had a significant impact on the mental health of healthcare workers. (1–14) According to the World Innovation Summit for Health, among healthcare workers anxiety symptoms ranged between 23% and 46%, depression symptoms between 21% and 37%; and post-traumatic stress symptoms ranged between 17% and 50% for those who worked in COVID-19 units.(15) Despite the overwhelming evidence on the impact of COVID-19 pandemic on the mental health of health workers, evidence is comparatively scarce for South America. The HEROES study is one of the few large-scale surveys exploring this issue and found that in Latin America, Chile had the highest rate of health care workers with depressive symptoms (36.7%), symptoms consistent with severe depression (15.4%), and suicidal ideation (15%).(16)

Besides the high rates of mental health problems during the COVID-19 pandemic, Chile has some particularities that makes it interesting to study further. Due to its temperate climate, respiratory diseases are highly seasonal, concentrated in the winter, which means the health service suspends elective procedures to convert wards into basic and advanced respiratory units and increase hospital and intensive care capacity temporally during the winter months. (17, 18) Additionally, Chile had a high number of cases and admissions due to H1N1 Influenza during the pandemic in 2009,(19) leading to an increase in hospital capacity to deal with the emergency. Both factors could translate into efficient plans for dealing with a respiratory disease emergency, and, therefore, the experience of healthcare workers might be different to what has been reported elsewhere. Therefore, this study explored how intensive care unit healthcare workers experienced their hospital response prior to and during the COVID-19 pandemic in seven centres in Chile. Exploring the perspectives of healthcare workers caring for the most severe patients could help us learn from how the response was deployed to improve preparation for future epidemics or catastrophes that require increasing acute care hospital capacity quickly.

## Methods

We adopted an exploratory qualitative methodology, conducting interviews with healthcare professionals who had worked in an intensive care unit during the COVID-19 pandemic in Chile. In our protocol,(20) we planned to conduct focus groups organised by profession, but after four failed attempts (i.e., none of the scheduled participants attended), we decided to change data collection to 1:1 interviews. The reporting of this study follows the standards for reporting qualitative research.(21) (See S1 File)

The topic guide focused on three areas:

1. Functions within and relationship with the team: exploring whether the interviewee felt part of the team and their relationship with colleagues and their line manager.
2. Organisational preparation: activities conducted within their organisation/ team to prepare for and in response to the COVID-19 pandemic. This includes procurement of new staff, contingency plans, adaptations to increase bed capacity, and psychological support.
3. Cognitive, physical, and organisational challenges: referring to the experience of the team during the pandemic, in particular, how they dealt with uncertainty and how new staff were integrated into the team to provide patient care without compromising quality of care.

We conducted semi-structured interviews between November 2020 and March 2022 over the telephone or via videocall, lasting between 30 minutes to one hour. A clinical psychologist trained in qualitative methods acted as an external mediator, guided the discussion and provided emotional support if or when the interview topics were too sensitive. The researchers were not involved in conducting the interviews and had no personal relationship with the interviewees.

Ethical approval was granted by the Clínica Alemana Research and Clinical Trials Unit, Faculty of Medicine, Clínica Alemana Universidad del Desarrollo Ethics Committee (registration number 2020–78) and the Servicio de Salud Metropolitano Oriente Ethics Committee (registration number 152–0029) before recruitment began. Data collection started on the 24^th^ of November 2020 and ended on the 19^th^ of March 2022.

### Context when the study was conducted

Chile identified their first COVID-19 case on the 3^rd^ of March 2020. As a measure to contain the spread of the virus, the government closed schools and universities on the 15^th^ of march, while social gatherings were prohibited, and borders were closed on the 18^th^ of March.(22) Chile initially implemented selective lockdowns, with the whole capital and some adjacent local authorities entering lockdown on the 15^th^ of May, when 78% of the ICU beds were occupied.(23–25) The use of facemasks was mandatory in public spaces (indoors or outdoors) and people were only allowed to leave their home for specific activities, such as grocery shopping, as long as they had a virtual permit issued by the police. Additionally, the private and public sector hospitals were brought together under a single administrative unit led by the ministry of health. This meant that inpatient and intensive care beds could be allocated centrally, regardless of where the admission happened.(26)

On the 19^th^ of July 2020,(27) the government decided to implement a plan for gradually opening services to the public. This plan consisted in five different levels of restrictions by local authority depending on the number of new cases detected per week. The highest level of restriction was complete lockdown. The following level (i.e. transition) capped the number of people that could gather indoor at 1 person per 4 m2, there was a curfew between 11 PM and 5 AM, and people could not go through or visit local authorities in lockdown.(28) The use of facemasks remained mandatory until October 1^st^, 2022.(29)

### Sampling and recruitment

We purposively sampled nurses, physiotherapists, healthcare assistants and consultants (ICU or other specialty) who worked in an intensive care unit in seven centres participating in the IMPACCT COVID-19 study. We sampled for variation in gender, profession, geographical location and years of experience in ICU. Participants were recruited through posters in their workplaces and reminders were sent via their clinical WhatsApp group. Potential participants were asked to complete a brief survey collecting their contact details, basic demographics and potential times/dates when they could meet for the interview. The participant information sheet and consent form were attached to this brief survey. Verbal consent was obtained before commencing the interview and any queries they might have had were answered.

### Data analysis

Our analytical approach was inductive without following a predefined theory. We used transcriptions, interview notes and recordings to analyse data thematically. Two researchers independently coded the interviews following the steps of framework analysis.(30) Weekly meetings were held to enable triangulation and constant updating and refining of the coding framework. After all the interviews had been analysed, we created a summary of the findings and our interpretation, which were shared with an independent sample of professionals in a focus group (FG). The FG comprised five participants, two physiotherapists, and three nurses who were invited via WhatsApp. These participants completed the same brief survey as the participants who were interviewed. The FG followed a topic guide (see S2 File) with open questions and was conducted virtually using the Zoom platform. The quotes used are presented in the results section using FG as part of the anonymization code. The initial coding framework was further refined after including the discussion from the focus group.

### Reflexivity

Throughout the design and analysis phases, we maintained a reflexive approach to prevent the influence of our preconceptions on the research findings. All the researchers were female, physiotherapists, and have experience working in intensive care; it is possible that our experiences may have influenced our interpretations of women healthcare professionals. ACA was a frontline clinician during the swine flu pandemic in 2009. To avoid the impact of these potential biases, the data collection was conducted by an independent clinical psychologist, and we triangulated our data analysis in two ways: two researchers analysed the data independently and initial findings were shared with a different sample of eligible professionals. This allowed us to sense check our interpretations. We also held monthly meeting with the wider research team to gain deeper understanding of what was happening on the ground.

## Results

Interviews with 38 healthcare professionals, lasting between 22 and 54 minutes, were conducted between November 2020 and March 2022. We interviewed more women than men (23/38), 15 (40%) participants were physiotherapists, and 21 (56%) professionals had between 3 and 10 years of experience working in ICU (see Table 1).

**Table 1.** Characteristics of participants.

| Characteristics | n=38 |
| --- | --- |
| Gender |  |
| - Female | 23 (61%%) |
| - Male | 15 (39%) |
| Profession |  |
| - Physiotherapist | 15 (40%) |
| - Nurse | 9 (24%) |
| - ICU Consultant | 7 (18%) |
| - Healthcare assistant | 7 (18%) |
| Years of experience at ICU |  |
| - Less than 3 years | 10 (26%) |
| - Between 3 and 10 years | 21 (56%) |
| - More than 10 years | 7 (18%) |

### Measures implemented to increase bed capacity

All centres increased intensive care bed capacity by converting other units into ICUs, relocating ICU staff to the new units, and training staff from other non-ICU wards. Additionally, in some centres new staff were hired at the beginning of the first wave (i.e., April 2020), while in others this happened much later. Table 2 shows in detail the changes implemented to increased staff in each participating centre.

**Table 2.** Summary measures adopted by each centre to increase hospital capacity.

|  | Staff | Bed capacity | Working hours pattern | Human resource training | Mechanical ventilators |
| --- | --- | --- | --- | --- | --- |
| Hospital_1 | Hiring of agency staff<br>Transfer of staff from other units to the ICU (Nurse_1/Consultant_3)<br>New agency healthcare assistants (HA_1) | Increase of ICU beds from 12 to 64 (Nurse_5)<br>Conversion of other units into ICUs (Consultant_3) | 24 hours working: three free days (Physio_11/ Nurse_1/ Nurse_5/ HA_1)<br>Many doubled their shifts, and the residents did up to three shifts per week (Consultant_3) | Online training on how to use mechanical ventilators (Nurse_1)<br>Training sessions were very basic. Someone would come over and explain to whoever was free. The session was recorded on a phone, so it could be shared with other colleagues (Physio_11) | We used to have three types of MV and now we have ten different ones (Physio_11)<br>We increased the number of mechanical ventilators (Nurse_1 /Consultant_3) |
| Hospital_2 | Increase of agency staff (Nurse_3/ Physio_5 / Consultant_4) | Progressive opening of new beds depending on need, from six ICU beds up to 47 (Physio_5/ Physio_9 /Consultant_4)<br>20 more ICU beds and an entire floor were opened<br>Conversion of units into ICU (Nurse_3 /Nurse_2) | 24 hours working: three free days (Nurse_3/ Physio_5/ Physio_9)<br>Mandatory extra shifts (Consultant_4) | No formal training sessions (Nurse_3/ Nurse_2 / Physio_9) | Increase in the number of ventilators and respiratory equipment (Nurse_3/ Nurse_2/ Physio_9) |
| Hospital_3 | Professionals from other units working in the ICU (HA_3)<br>Hiring of agency staff (Physio_1/ Physio_4/ Physio_7) | Increase more than double in ICU beds (Physio_8 / HA_3/ Physio_1) | 24 hours working: three free days (Physio_1/ Physio_4/ Physio_7/ Physio_8/ Nurse_4/ HA_3) | Training on the use of new ventilators (Physio_8), and use of PPE (Physio_1/ Physio_4/ Physio_7) | Increase in the number of ventilators (Physio_1/ Physio_7/ Physio_8/ Nurse_4/ HA_3) |
| Hospital_4 | New staff were paid as freelance (Physio_2)<br>Physiotherapy staff were doubled (Physio_6)<br>New staff hired were inexperienced (Nurse_7) | Increase in bed capacity to reach 133 (Nurse_7 / HA_5/ Consultant_5)<br>New hospital wards were transformed into ICU | 24 hours working: three free days (Physio_2 / Nurse_7 / Nurse_9/ HA_5) | Training on the use of mechanical ventilators (Nurse_7 / Nurse_9)<br>Training on the use of PPE and flow of COVID patients | Mechanical ventilators were sent by the government, and some were borrowed from a contractor (Kines_2/ Physio_6/ Nurse_7/ |
|  | Staff were transferred from different units to the ICUs (Nurse_9)<br>Agency staff were hired during the 2nd wave (Consultant_6) | beds (Nurse_9/<br>Consultant_5) | Weekly changes of shifts (Consultant_5/<br>Consultant_6) | (Physio_6) | Nurse_9 / HA_5/<br>Consultant_5/<br>Consultant_6) |
| Hospital_5 | Staff were transferred from different units to the ICUs (Physio_3 / Consultant_1)<br>During the second wave, the staff tripled (Physio_3) | Bed increase and conversion of units (Nurse_8/ Physio_3/<br>HA_4/ HA_6/<br>Consultant_2) | Working hours remained the same (Nurse_8/<br>Physio_3), but consultants had more shifts per week (Consultant_1) | There were no formal training sessions (Physio_3/ Consultant_2) | New mechanical ventilators (Physio_3/<br>HA_6/ Consultant_1) and medical supplies (HA_4/ Consultant_2) were purchased |
| Hospital_6 | There was an increase in physiotherapy staffing to reach a staff: patient ratio of 1:6 (Physio_10/ Physio_13)<br>That meant, we were five initially and increased to 12 physios (Physio_10) | Initially, ICU beds were increased from 8 to 20 (Physio_13/ HA_2/<br>HA_7)<br>Other wards were transformed into ICU (Nurse_6/ Physio_10)<br>To create 40 extra ICU beds (Physio_13) | 24 hours working: three free days (Physio_10/<br>Physio_13/ HA_2) | We received training to use the new devices (Physio_10)<br>Also, training regarding the use of PPE (Physio_13)<br>We received training to move patient in bed and the use of PPE (HA_2) | Increase in the number of invasive and non-invasive ventilators and nasal high flow (Physio_10/<br>Physio_13)<br>Ventilators increase (HA_2) |
| Hospital_7 | Staff increase (Physio_12) | Increase in beds and conversion of units (Physio_12) | 24 hours working: three free days (Physio_12) | Training on the use of EPP and use of new ventilators (Physio_12) | Increase in the number of ventilators (Physio_12) |

159 Physio: Physiotherapist; HA: Healthcare assistant; Consultant: ICU consultant; ICU: intensive care unit; PPE: personal protective equipment.

The changes that occurred in the short term meant increased tiredness, greater susceptibility, and a lower emotional threshold to different work situations, such as the death of patients, which generated anger, stress, a sense of powerlessness. This greater emotional liability was paired with increased workload and changes in working shifts. Several workers had to resort to private psychological support since the support offer from their institutions was insufficient.

In terms of the experience of these professionals working in the ICU, we identified seven themes:

### “As the pandemic was unfolding”

The preparation before the arrival of COVID-19 in Chile varied in the different participating sites, but as the pandemic was unfolding new units were converted and opened.

> “The whole 2020 was very traumatic. There was no more human resource, but they still opened more beds and units” Physio_3

> “At some point, we were moved out of our units to install a new oxygen network, so we could have mechanical ventilators. In that time, when we went back to our units, everything was ready. We were told ‘We will be an ICU tomorrow’ and that’s how we had to learn as it happened” Nurse_1

> “As more beds were needed, and the ICU was full, HDU beds were converted into ventilated beds. Everything was being converted depending on need” Consultant_2

> “I remember that they took a long time to give instructions from the top. We already knew […] we were going to be converted. We were willing to do all the changes. We were worried about what was coming. When it became evident [the pandemic], I remember that the leadership took quite a while to make decisions” Consultant_3

Clinical staff was transferred from different units to work on the different ICUs and new converted units.

> “In the ICU physiotherapy team, we were very few. Later, the colleagues that worked in the outpatient physiotherapy service became intensive care physios” Physio_4

> “I used to work in another place that was musculoskeletal. Just before the pandemic started, I began working in HDU, that’s how I ended up in the ICU” Physio_2

The staff that were hired, in general, were recently graduated healthcare professionals or without the required experience to work in ICU. Many of these professionals were agency staff or worked as freelance.

In one particular case, Physio_3 said there was no increase in the number of staff until the third wave (i.e., October 2021); however, when it increased, it was a threefold increase.

Only two hospitals of the seven participating sites prepared in advance regarding physiotherapy staffing levels (Physio_2, Physio_6, and Physio_10), however it is noteworthy that in both institutions, the change to the physiotherapy staffing levels was being planned since before the pandemic.

In all the participating sites, the increase in ICU beds was organised depending on the demands of the pandemic, starting in March 2020, and was adapted in line with the government requirements.

Training for using new equipment, as well as personal protective equipment (PPE), high flow nasal cannulas (HFHC), invasive and non-invasive mechanical ventilators were conducted, however, most interviewees mentioned that these training sessions were not appropriate because staff members did not have time to attend or the training materials were sent as short clips. On some occasions, training was delivered too late (e.g., the equipment was already being used) or simple it was never delivered.

> “They told us we were going to be trained, but actually, we were sent short clips. Nobody showed up to train us” Nurse_1

In most cases, the training sessions were insufficient for the new members of staff, as these were quick and very general.

In addition, Nurse_7 highlighted that, due to the public health emergency context, it was not always possible that all members of staff attended training sessions, therefore, as a way of helping each other, the colleagues that were able to attend would feedback the information to the rest of the team. Similarly, Nurse_9 said that in her hospital everyone was summoned to attend the training sessions every time new equipment arrived.

> “I learned as the pandemic was evolving. Listening and listening to the teachers when I was a resident, and we tried to do our best” Consultant _3

> “It was the first time I was in an ICU, because this was my first job and had never been in an ICU. [Everyone was] super nice to teach me everything from the beginning because what one was learning was whatever one was seeing. I didn’t know about intubations, I didn’t know anything, and bit by bit, they taught me” HA_6

### Change of usual roles

To ameliorate the cultural and organisational changes that suddenly happened with the addition of new members of staff and the opening of new beds, workers were allocated to different roles, for example, as shift managers, in mixed teams with experienced and inexperienced staff, and assigned to a different shift rota.

The rationale behind these changes was that many of the new members did not have experience working in ICU or were not used to working with critically ill patients. Staff members with more experience were allocated to the new units, assuming roles as tutors and supervisors while performing their usual work. This led to additional stress and, in many cases, the loss of existing networks of social support with the colleagues sharing the same rota.

> The staff had to be divided. Some of them went into the clean ICU and others stayed in the COVID ICU. Therefore, the staff from the HDU had to be trained, but they didn’t have enough expertise, so we had to be like their older sibling and care for the most severe patients” Nurse_8

> “We received agency staff, and later, recently qualified colleagues, which was quite complex for us because, for example, I was the shift supervisor and on top of that, I had to take care of the patients that were assigned to the new nurses, because they didn’t have the basic skills” Nurse_3

> “The ICU consultants and the physios created technical managers. That was something new. The colleagues that moved into that model were the most experienced ones. They moved into the day shifts and made sure the decisions were the most appropriate ones” Physio_11

> “People who didn’t work in ICU, those from the medical-surgical [unit], the OR, in general, everyone moved to the ICU, because their units had to be converted, too” Physio_14

> “Staff who worked in ICU was distributed to other units so they can provide support or supervision, so there’s could be continuity and also to try to homogenise with those with less experience” Consultant _1

> “All the doctors doing an internal medicine residency… that is, 1st, 2nd, and 3rd year. All of them had clinical duties assigned, instead of academic ones, and started to take responsibilities that were not on a par with their training” Consultant_3

### Social isolation, exhaustion, and lack of psychological support

Emotional regulation was a challenge many healthcare professionals had to face during the COVID-19 pandemic. For some professionals, their personal life supports their emotional regulation, while for others, the organisational culture is the main source of support, because their professional image is considered a key aspect of their work life.

The professionals who lived with their families, had to isolate or distance themselves to avoid spreading the virus. This meant leaving their homes, implementing hospital-grade disinfection protocols or avoiding direct contact with their family members. Additionally, the focus group participants highlighted the extra costs they had to cover to keep their families safe, which put them into economic hardship. This was compounded by the lower salaries permanent staff had compared with agency staff, which made permanent staff feel undervalued.

> “I had to move houses because I was living with my parents. So, I couldn’t keep living with them with COVID [going around] because both of them are high-risk. I had no other option, but to leave” HA_1

> “We left a lot of things behind for work. Family, partner, friends, we even stopped having a social life… and nobody cared about us” HA_2

> “Fear, fatigue, frustration, and a lot of anger. There was plenty of anger. Sometimes I would think ‘Why do I have to sacrifice myself?’… when the rest of the population are not even wearing masks. Which, in a way, is a very human response to exhaustion” Consultant _5

The emotional burden plus the organisational changes implemented to meet the healthcare demands created a negative environment and culture with lack of support and increased workload, which led to multiple deleterious effects such as panic attacks, emotional instability, frustration, anger, and exhaustion that, on many occasions, required sick leave. This mix of factors contributed to increased physical and emotional exhaustion.

> “The hospital director did not appreciate us; he always demanded too much as he found our job was not worth it enough. It was just about opening more beds, but they did not see the healthcarés quality we have been providing” FG _Nurse 1

> “It was quite tough, with a significant increase in beds and a lot more patients. We were all putting in more time than usual; everyone was asked to take extra shifts. It was quite challenging, since some of us started taking leave, and many were dealing with depression” Consultant _4

The increase in sick leave led to greater workloads because the professionals had to cover their rota and additional shifts to cover for their colleagues on sick leave. The organisational measures to provide psychological support for the stress, exhaustion and possible burnout staff were experiencing were deemed as insufficient, late or inappropriate.

> “They provided psychological support for people who felt overwhelmed… one could go spontaneously and meet with a psychologist to express what was happening or what they were feeling. There was not a more effective approach; instead, our team had to cope with it and go out to find the appropriate therapist” Consultant _5

> “I personally remember that the staff wellbeing office began broadcasting uplifting messages over the loudspeakers, hoping that everyone would be well. However, it was actually frustrating because instead of helping, it overwhelmed the entire team” Consultant _6

The support provided by their organisations was not evident for most. Some interviewees mentioned that the process to access support was cumbersome or inaccessible, and at times, they did not perceive any improvements in the access to psychological support even when their line manager was aware of the situation.

### Fear of becoming a vector for infection

The professionals created hospital-grade disinfection protocols in their homes and with their families due to the fear of becoming a vector of infection. Many of our interviewees mentioned that they stopped visiting their elderly parents or family members for months to avoid exposing them to the virus.

> “I had young children I wanted to hug once I arrived home… Nevertheless, I would come home directly to take a shower, and put my clothes straight into the washing machine after disinfection. Therefore, it was a whole ritual before hugging my kids” Nurse_7

> “I live with my mum, and I had to move out to protect her from the disease. We were all full of fear. I had to stay with a friend who also worked in the same field to avoid infecting mum” Physio _1

### “Keep calm and carry on” mentality

Seventeen percent of our interviewees referred to have requested sick leave due to mental health issues. Those that continued working felt that they “had to continue anyway” because there was no staff to cover the rota. The healthcare professionals unconsciously made personal sacrifices to meet the demand of the COVID-19 pandemic, which for some of our interviewees worsen the situation and led to repeated and long mental health sick leave.

> “Extra shifts were a must. There were not enough people …which made it extremely challenging to manage the shifts. The workload increased significantly and unexpectedly… I worked so many hours that I would stay in the hospital, studying or reading, until the next shift. That was, in fact, so intense that it felt very depersonalized… I never stopped working; it was like being in a treadmill, and I was not aware what I was doing to myself” Physio_12

> “At first, the team just went for it, and I believe nobody questioned this… there is a very high emotional cost; many of us ended up extremely exhausted” Consultant _7

> “We always repeated to ourselves ‘Well, this is it. We have to do the best we can and move forward’” Nurse_5

### “Depersonalisation”: balancing doing their job and keeping emotions under control

The combination of a high volume of patients, high turnover, and the increasing demand for beds meant that there was no time to grieve when a patient died. Additionally, because visitors were not allowed inside the units, on many occasions, the healthcare professionals would spend time next to a dying patient to keep them company. The tacit expectation to remain stoic, “do the job and keep going” together with the number of patients dying daily, meant that they had to ignore what they felt and keep distance with their emotions. Many described that they had lost the human touch and treating patients was similar to being in an assembly line.

ICU consultants mentioned that it was difficult “to keep it together” when they had a patient dying alone in their unit, and a grieving family on the phone that could not see what was going on but still needed emotional support.

> “Depersonalisation happened a lot. Someone would pass away, and it was just a matter of cleaning up. It was like, let’s clean up, and another patient. The patient arrived, and the family was not there, which was terrible… We just had to keep going. When everything settled in, I could feel it. It hurts to remember how tough it was. My voice breaks” FG_Nurse_3

> “I believe the hardest part was having young, vital, and functional patients with their families… plenty of them on ventilators passed away, and having to support the family. Even though we felt broken, obviously, that could not be demonstrated” Consultant _3

### Uncertainty

Interviewees mentioned that when they first received patients with COVID-19, their unit, line managers and themselves did not know how to treat and care for these patients. Uncertainty was omnipresent. From what personal protective equipment they should wear to what parameters they should use to ventilate these patients. This meant that health professionals learnt by trial and error and had to constantly adapt as more evidence was emerging from other countries.

> “At the beginning of the pandemic, we had very imprecise notions of this disease. We actually knew nothing… there was imprecise information about patient admission flows and what we were going to do to assist them” Consultant _5

> “Never will I forget. Nobody wanted to be the first treating these patients… There was a lot of fear, denial, uncertainty as the clinical team did not know whether we were going to treat them properly” HA_4

## Discussion

Our multicentre, qualitative interview study focusing on the experience of healthcare workers in the intensive care unit had similar findings to the available literature, highlighting that, despite different policy responses at the country, regional and local level, the experience of front-line workers was comparable. Similar to other studies, the pandemic response was perceived as constantly changing(5, 6, 9) (theme 1) and adapting to the situation, which led to changes in the way of working to cope with the lack of trained staff(2, 5, 11, 31) (theme 2).

This everchanging environment was partly due to high levels of uncertainty (theme 7) on what the best course of treatment, but also facing unfamiliar ethical and emotional challenges (e.g., restrictive visiting policies, end-of-life care).(11–13, 32) At a personal level, our participants had to juggle protecting their love ones and maintaining a family life (theme 4), which was a common worry among health care workers during the COVID-19 pandemic.(9–11, 14) Similar to the findings by Rao et al (2021)(12), our participants accepted the situation as part of their professional responsibility (theme 5); however, in the long run, they experienced emotional distress, sadness, anger, and exhaustion (theme 3), which again echoes the findings from other countries.(2–4, 8, 11–13, 31) The combination of these experiences might have led to depersonalisation (theme 6), where healthcare workers suffer from compassion fatigue as a protective mechanism, but also as a red flag of burnout symptoms.(3, 12, 14, 32)

The declaration of the pandemic in March 2020 and the lockdown measures imposed due to the SARS-CoV-2 outbreak generated an unprecedented emergency response, with consequences for the mental health of the general population and especially healthcare workers. Several systematic reviews report an increase in the prevalence of stress, anxiety, depression, insomnia, or burnout among healthcare workers during the COVID-19 pandemic.(2, 10, 33, 34) The impact on the mental health of healthcare workers had already been documented during the outbreaks of SARS-CoV-1 (2003), H1N1 (2009), MERS-CoV (2012), or Ebola (2014), showing moderate and high levels of anxiety, depression, post-traumatic stress, and absenteeism. (34–36).

One previous study(37) suggest that stress-related psychosomatic symptoms in healthcare workers are associated with the social pressure they experience, where society expects them to address problems relentlessly, even when these problems are out of their control. This leads to depersonalisation, stress, job dissatisfaction, and burnout symptoms.(37) The stress, emotional exhaustion, and depersonalisation described by our interviewees could be seen as signs of burnout syndrome, which was frequent among Chilean ICU healthcare workers during the COVID-19 pandemic, ranging between 24.5% and 46.1%.(38) This is relevant because having burnout can increase the risk of making avoidable mistakes with patients by 60%.(37)

Salehi et al. [33] suggest that job satisfaction is a key factor for retaining staff during health crises.(39) Factors such as job security, time flexibility, role transparency and participation in decision-making increase the willingness of staff to stay in an organisation, reducing job burnout and turnover.(39) On the other hand, availability of PPE, adequate rest, and a safe environment were linked to lower job dissatisfaction and better quality of care. (39) Although our participants did not refer directly to their job satisfaction, their experience of working in a high-pressure environment, constantly changing and where they felt exposed to a deadly virus could have long-lasting impact on their intention to leave the profession, and therefore, their experience should not be dismissed when planning the response to the next pandemic.

Improving retention and decreasing turnover of highly trained healthcare workers, such as those that work in the ICU, are key for the implementation of an early pandemic response. Evidence from the SARS and MERS pandemics shows that the main limitation to expand intensive care capacity was lack of nursing staff with ICU experience, which cannot be easily procured once the pandemic has begun.(10)

Interventions and strategies that that the literature suggest based on lessons learned from the COVID-19, SARS and MERS resonate with some of the problems our participants described. Uncertainty. confusion, lack and overload of information are expected in an emergency, but this should be acknowledged.(8, 10, 14) Having a leader or “champion” has been proposed as a potential solution to communication issues because they can foster a sense of coherence,(10) provide clear and regular information about planning and policy changes,(8, 14) and engage frontline workers in decision-making.(8, 10, 13) Lastly, having plans in place to expand capacity in terms of PPE, recruitment of new staff including their training and support, and procurement of skilled staff are key to maximise the wellbeing of healthcare workers.(8)

According to the HEROES study, Chile is the country that presented the highest number of cases with suspected depressive symptoms and suicidal ideation. For this reason, is essential to protect the mental health of workers by developing programs that provide psychological and medical support to those who need it. Factors associated with worse mental health during the COVID-19 pandemic, which were also mentioned by our participants are: lack of social and economic support, concerns of infecting family members, increased workload, and change of roles and functions in the workplace.(16, 40)

### Limitations

This study explored the healthcare workers’ perceptions and experiences of the changes they faced while working in an intensive care unit during the COVID-19 pandemic. Although we extended the invitation to participate to all professionals working in the participating centres and we attempted to have equal representation from the different professions, it is possible those who agreed to the interview had an interest in sharing their opinion because they had a bad experience or were in leadership positions, and therefore, we did not capture the opinions of those who had a more positive experience.

## Conclusion

The healthcare workers’ experiences and perceptions of working in ICU during the COVID-19 pandemic are the result of the psychological effects of the restrictions imposed in the country, a high volume and workload, limited preparation at their hospitals, and fast changing environment as the pandemic was unfolding. Because healthcare professionals are a scarce and valuable resource during public health emergencies, planning should be in place to rapidly increase ICU staff in the event of a new prolonged emergency without depleting existing human resources. Appropriate psychological and emotional support should be provided in a timely fashion.

## Acknowledgements

We would like to thank Cristián Rosales-Antequera for his help with earlier versions of this manuscript.

## Study registration

NCT04979897

## Data statement

Due to the sensitive nature of the questions asked in this study, participants were assured raw data would remain confidential and would not be shared.

